# The causal associations between cancers and cardiovascular diseases: a two-sample bidirectional Mendelian randomization analysis

**DOI:** 10.1101/2023.09.18.23295757

**Authors:** Daoming Zhang, Haoyue Li, Yuan Li, Tian Tang, Zhenming Fu, Yongfa Zheng, Ximing Xu

**Author notes:** Correspondence to: Zhenming Fu, MD; Yongfa Zheng, MD, and Ximing Xu, MD, Cancer Center, Renmin Hospital of Wuhan University 238 Jiefang Road, Wuhan, Hubei Province 430060, China. D. Zhang, H. Li and Y Li contributed equally.

## Abstract

**BACKGROUND:** Associations between cancer and cardiovascular disease (CVD) have been reported previously in observational studies. However, the causal relationship between the specific subspecies of the two diseases remains unclear. This study used a two-sample bidirectional MR study to investigate the causal relationship between different types of CVDs and the major types of malignancies and vice-versa.

**METHODS AND RESULTS:** We extracted summary statistics for coronary atherosclerosis, hypertension, hypertrophic cardiomyopathy, heart failure, atrial fibrillation, stroke, and 14 common malignancies from published relevant genome-wide association studies as instrumental variables. We conducted two-sample bidirectional Mendelian randomization (MR) studies to assess the causal relationship between CVD and cancer in which the inverse variance weighting (IVW) method was the main method. Multiple comparison calibration, sensitivity analysis, and heterogeneity analysis were performed to improve the reliability and robustness of the results. The evidence from IVW analyses showed that genetically predicted coronary atherosclerosis was suggestively associated with a decreased risk of endometrial cancer (OR=0.053, 95% CI: 0.004-0.648, P=0.022); hypertension was suggestively associated with an increased risk of oral cavity/pharyngeal cancer (OR=14.872, 95% CI: 1.324-167.053, P=0.029); hypertrophic cardiomyopathy was suggestively associated with a decreased risk of brain cancer (OR=0.479, 95% CI: 0.257-0.890, P=0.020); any stroke was suggestively associated with a decreased risk of breast cancer (OR=0.798, 95% CI: 0.669-0.952, P=0.012) and prostate cancer (OR=0.844, 95% CI: 0.737-0.966, P=0.014) since their significance weakened after multiple testing. In the reverse MR analysis, bladder cancer was associated with an increased risk of coronary atherosclerosis (OR=1.426, 95% CI: 1.051-1.934, P= 0.023) and hypertension (OR=1.689, 95% CI: 1.115-2.557, P=0.013); pancreatic cancer was associated with an increased risk of any stroke (OR= 1.047, 95% CI: 1.005-1.090, P= 0.027), losing significance after multivariate testing. Prostate cancer was significantly associated with an increased risk of heart failure (OR= 1.030, 95% CI: 1.009-1.053, P= 0.006); cervical cancer was significantly associated with an increased risk of any stroke (OR= 8.751686e+03, 95% CI: 35.043-2.185650e+06, P= 0.001).

**CONCLUSIONS:** Causal relationships for specific types of CVD and cancer were found in this MR Study, although some were suggestive. This study provides ideas for the follow-up management of these two common chronic diseases.

**CLINICAL PERSPECTIVE:** *What Is New?:*

- Some observational studies have shown that cardiovascular diseases (CVDs) and cancer have complex causal relationships dominated by positive associations. However, the role of genetic factors in their comorbidities remains unclear.
- In this study, by utilizing data from genome-wide association studies, we identified a significant genetic correlation between multiple groups of specific classes of CVDs and specific types of malignancy, along with the shared risk snp. Some of the results are contrary to previous reports and warrant further research. These findings could provide insights into the shared genetic architecture between CVD and cancer.

*What Are the Clinical Implications?:* This study adds to the understanding of the underlying causal relationships of different phenotypes of CVD and cancer, with implications for the prediction and prevention of these common comorbidities.

---

Cardiovascular disease (CVD) and cancer are common diseases worldwide, seriously affecting survival time and quality of life. Malignant tumor is the second leading cause of death globally. In 2020, 19.29 million new cancer cases worldwide were reported, with 9.96 million deaths. With an aging global population, the cancer burden is expected to increase by 47% in 2040 compared to 2020.^1^ Cancer threatens human health and poor quality of life. The risk factors are highly complex and heterogeneous. Cancer prevention focuses on specific risk factors such as tobacco use, diet, lifestyle habits, and carcinogen infections.^2-4^ CVDs are the leading cause of the global disease burden. The number of deaths attributed to CVD increased by 18.7% from 2010 to 2020.^5^ Currently recognized common risk factors for CVDs include hypertension, hyperlipidemia, obesity, diabetes, smoking, and alcohol consumption.^6^ The coexistence or sequential occurrence of the two diseases is common. The intricate potential causal relationship between them has long attracted the interest of researchers and related epidemiological studies have been reported; however, these have focused on specific types of studies in two categories of diseases.

In a prospective follow-up study of individuals who did not have cancer at baseline, the prevalence of CVDs and occurrence of interim cardiovascular events were not associated with a higher risk for subsequent cancer development. However, ideal cardiovascular fitness is associated with a lower risk of future cancer. Studies have shown the association between CVD and future cancer which is attributed to shared risk factors.^7^ An observational study of long-term CVD risk in 1,26,120 cancer survivors suggests that cancer therapies, particularly chemotherapy, rather than cancer itself, play a major role in CVD risk.^8^ In contrast, in a Canadian study, newly diagnosed cancer was associated with higher risks for cardiovascular mortality, acute myocardial infarction, stroke, and heart failure after adjustment for covariates including age, sex, hypertension, diabetes, dyslipidemia, and prior CVD.^9^ Possible explanations for the conflicting results to date include the degree to which some studies were effective in adjusting for shared risk factors and the vulnerability of previous studies to biases inherent in observational studies, such as residual confounding and reverse causality. Simultaneously, the intricate associations between different types of CVDs and malignancies cannot be studied generally without a breakdown.

Mendelian randomization (MR) estimates the causal relationship between exposure and outcome by using instrumental variables (IVs) strongly associated with exposure and strengthens the causal inference by controlling for non-genetic environmental confounders and reverse causality. MR analysis, due to random combinations of alleles during meiosis, and germ cell line genetic variation fixed at conception, is less susceptible to conventional confounders and reverse causation. Currently, no large-scale MR study examining the association between multiple CVDs and multiple malignancies has been reported.

This study used a two-sample bidirectional MR to investigate the causal relationship between different types of CVDs and the major types of malignancies and vice-versa.

## METHODS

### Study Design

A two-sample bidirectional MR study was designed to explore the impact of CVDs on the risk of common cancers as shown in **Figure 1**. The bidirectional MR study was based on three assumptions.^10^ All data were publicly available for both males and females and predominantly European population. The study adhered to the latest Strengthening the Reporting of Observational Studies in Epidemiology using Mendelian Randomisation (STROBE-MR) guidelines.^11^ The included studies were approved by the relevant ethical review boards, and informed consent was obtained from the participants.

**Figure 1.**
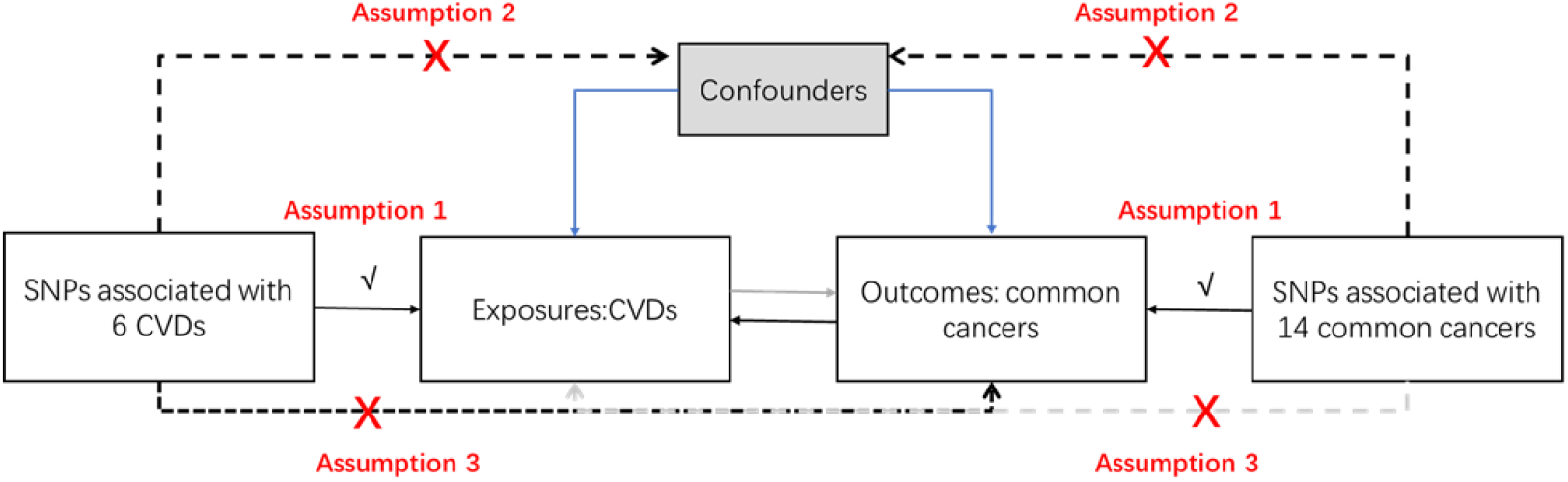
Study design. Dashed lines indicate a pleiotropic or direct causal relationship between exposure and outcome.

Single-nucleotide polymorphism (SNP) selection for CVDS was from genome-wide association studies (GWASs) of the UK Biobank, the FinnGen study, and other large consortia (**Table S1**). For CVDs, we detected six phenotypes including coronary atherosclerosis (361,194 participants with 14,334 cases and 346,860 controls), hypertension (463,010 participants with 54,358 cases and 408,652 controls),hypertrophic cardiomyopathy (337,159 participants with 71 cases and 337,088 controls), heart failure (977,323 participants with 47,309 cases and 930,014 controls), atrial fibrillation (463,010 participants with 5,669 cases and 457,341 controls), and any type of stroke (446,696 participants with 40,585 cases and 406,111 controls). Four criteria were used to select suitable SNPS as eligible IVs. First, selected SNPs were required to be suggestive of genome-wide significance (p < 5 × 10^-8^). Second, independence among the selected SNPs was assessed based on paired linkage disequilibrium. For r2<0.001 and clump window=10000-kb, independent SNPs without linkage were selected.^12^ SNPs with pleiotropic effects within or near the FTO gene were excluded.^13^ Third, F-statistics were calculated to verify the strength of individual SNPs. SNPs were considered strong enough to mitigate the effect of potential bias when the F-statistic was greater than 10. Fourth, to avoid potential pleiotropic effects, the PhenoScanner database was queried for secondary phenotypes for each SNP using a threshold of P < 1×10-5, and SNPs associated with outcome confounders were removed.^14^

### Common Data Sources of Cancer

We selected 14 specific types of tumors and their data sources and related information are presented in **Table S1**. The GWAS summary statistics of oral/pharyngeal cancer (4,671 participants with 2,342 cases and 2,329 controls) were obtained from the Oncoarray oral cavity and oropharyngeal cancer consortium.^15^ Thyroid cancer data (1,080 participants including 649 patients and 431 controls) were extracted from the report by Kohler et al.^16^ The GWAS summary data of brain cancer (33,832 participants including 15,748 patients and 18,084 controls) were obtained from Burrows et al. GWAS summary data for lung cancer (27,209 participants including 11,348 patients and 15,861 controls) were extracted from the International Lung Cancer Association (ILCCO).^17^ The GWAS summary data of breast cancer (33,832 participants including 15,748 patients and 18,084 controls) was obtained from the Breast Cancer Association Consortium (BCAC).^18^ Data on ovarian cancer (66,450 participants including 25,509 patients and 40,941 controls) were extracted from the Ovarian Cancer Associations Consortium (OCAC).^19^ Pancreatic cancer summary data (3,835 participants including 1,896 patients and 1,939 controls) were obtained from PanScan.^20^ Stomach cancer data were derived from the FinnGen study, including information on 633 patients and 218,159 controls. The GWAS summary data of liver cancer (372,184 participants including 168 patients and 372,016 controls) were obtained from the UK Biobank study. Data on colon cancer (361,194 participants including 2,226 patients and 358,968 controls) were extracted from the UK Biobank study. Data on endometrial cancer (121,885 participants including 12,906 patients and 108,979 controls) were obtained from a meta-GWAS published by O’Mara et al.^21^ The summary data on prostate cancer (140,254 participants including 79,148 patients and 61,106 controls) were from a GWAS published by Schumacher et al.^22^ Bladder cancer data were derived from a report by Burrows et al. which included information on 1,279 patients and 372,016 controls. The cervical cancer summary data included 563 patients and 198,523 controls. Since SNPs identified by GWAS in a proportion of cancers rarely reached the genome-wide significance level of P<5×10-8, we selected SNPs (P<1×10-5) as the IVs for these cancer species (**Table S2**).

### Statistical Analysis

Inverse-variance weighted (IVW) method was used as the main MR Method to estimate the interassociation. This approach calculates and combines Wald ratios for each SNP from meta-analyses to estimate the overall effect of exposure on outcomes.^23^ The MR-Egger, weighted median, and weighted mode analysis methods were used to supplement and increase the stability of the results. The MR-Egger method was used to assess the horizontal pleiotropy of selected IVs, When the Egger intercept of the linear regression was close to 0, there was no directional pleiotropy of the IVs, and the exclusivity hypothesis was considered valid.^24^ The weighted-median method can provide valid estimates if more than 50% of the information comes from valid IVs.^25^ The MR-Steiger method was analyzed to monitor the direction of potential causal effects between assigned exposures and outcome risks.

In terms of sensitivity analysis, Cochran’s Q-test was used to quantitatively analyze the magnitude of heterogeneity among genetic IVs, if the heterogeneity of the results was not significant (P>0.05), and the fixed effect model of IVW was used for MR analysis.^26^ MR Pleiotropy RESidual Sum and Outlier (MRPRESSO) and inverse variance weighted estimates were generated after the removal of outliers. The p-value of the MRPRESSO distortion test was used to assess significant differences between the estimates before and after outlier correction.^27^ Horizontal pleiotropy of SNPs was assessed using the MR Egger intercept method, if the intercept term was close to 0 (P< 0.05), and the effect of genetic pleiotropy was considered to be small. Leave-one-out sensitivity analysis was performed to test whether the results were influenced by any single SNP. For determining the exposure factors in the above meaningful relationships, the data in the corresponding FinnGen database were re-obtained, and MR analysis was performed. Meta-analysis was used to combine the association results from different databases. Correction for multiple testing was performed using the false discovery rate based on the BenjaminiHochberg method. All statistical tests were two-sided and were performed with the TwoSampleMR and MR-PRESSO packages in R Software 3.6.1.

### Ethics

As all studies included had been approved by their independent review boards, no additional ethical approval was required in our studies based on pooled curated data.

## RESULTS

### Screening SNPs

We screened SNPs associated with coronary atherosclerosis, hypertension, hypertrophic cardiomyopathy, heart failure, atrial fibrillation, and any other stroke using GWAS data. After querying the PhenoScanner database, some SNPs were eliminated due to associations with confounding factors for cancer occurrence, and SNPs associated with BMI and obesity were also excluded. Finally, the F-statistic >10 used to screen SNPs showed no weak instrument bias and were selected as IVs for MR analysis (**Table S2**).

### Causal Effects of Coronary Atherosclerosis on Common Cancers

The IVW method was used to analyze the effects of six types of CVDs on 14 common cancers. The results of MR analyses are presented in **Figure 2** and **Table S3**. The results showed that genetically predicted coronary atherosclerosis was associated with a decreased risk of endometrial cancer (OR=0.053, 95% CI: 0.004-0.648, P=0.022); hypertension was associated with an increased risk of oral cavity/pharyngeal cancer (OR=14.872, 95% CI: 1.324-167.053, P=0.029); hypertrophic cardiomyopathy was associated with a decreased risk of brain cancer (OR=0.479, 95% CI: 0.257-0.890, P=0.020); other strokes were associated with a decreased risk of breast cancer (OR=0.798, 95% CI: 0.669-0.952, P=0.012) and prostate cancer (OR=0.844, 95% CI: 0.737-0.966, P=0.014). Unfortunately, the significance of each causal relationship was attenuated after adjustment for multiple testing. After MR-PRESSO analysis for outlier detection, three outliers were identified in the association between coronary atherosclerosis and endometrial cancer (OR=0.095, 95% CI: 0.013-0.679, P=0.019), and one outlier was identified in the association between any stroke and breast cancer (OR=0.745, 95% CI: 0.652-0.853, P= 1.85E-05)(**Table S5**). The results of heterogeneity analyses and horizontal pleiotropy tests are shown in **Tables S4** and **S6**. For the association between hypertension and oral cavity/pharyngeal cancer, no heterogeneity among estimates of individual SNPs was detected in the Cochran Q-test (Q= 54.144; P= 0.545) and no pleiotropy was observed in the MR-Egger regression analysis (intercept P=0.875;), the same characteristic was applied to determine the association between any stroke and breast cancer. The scatter plots of the objects with statistically significant correlations (colored lines indicate the slopes of different regression analyses) are shown in **Figure S1**, and the forest plot of the results of the leave-one-out method is shown in **Figure S2**. MR-Steiger analysis was used to further verify the directionality of the above associations, and the results showed that the directions of causal associations between coronary atherosclerosis and endometrial cancer, hypertrophic cardiomyopathy and brain cancer, and other strokes, and prostate cancer were statistically significant (**Table S11**).

**Figure 2.**
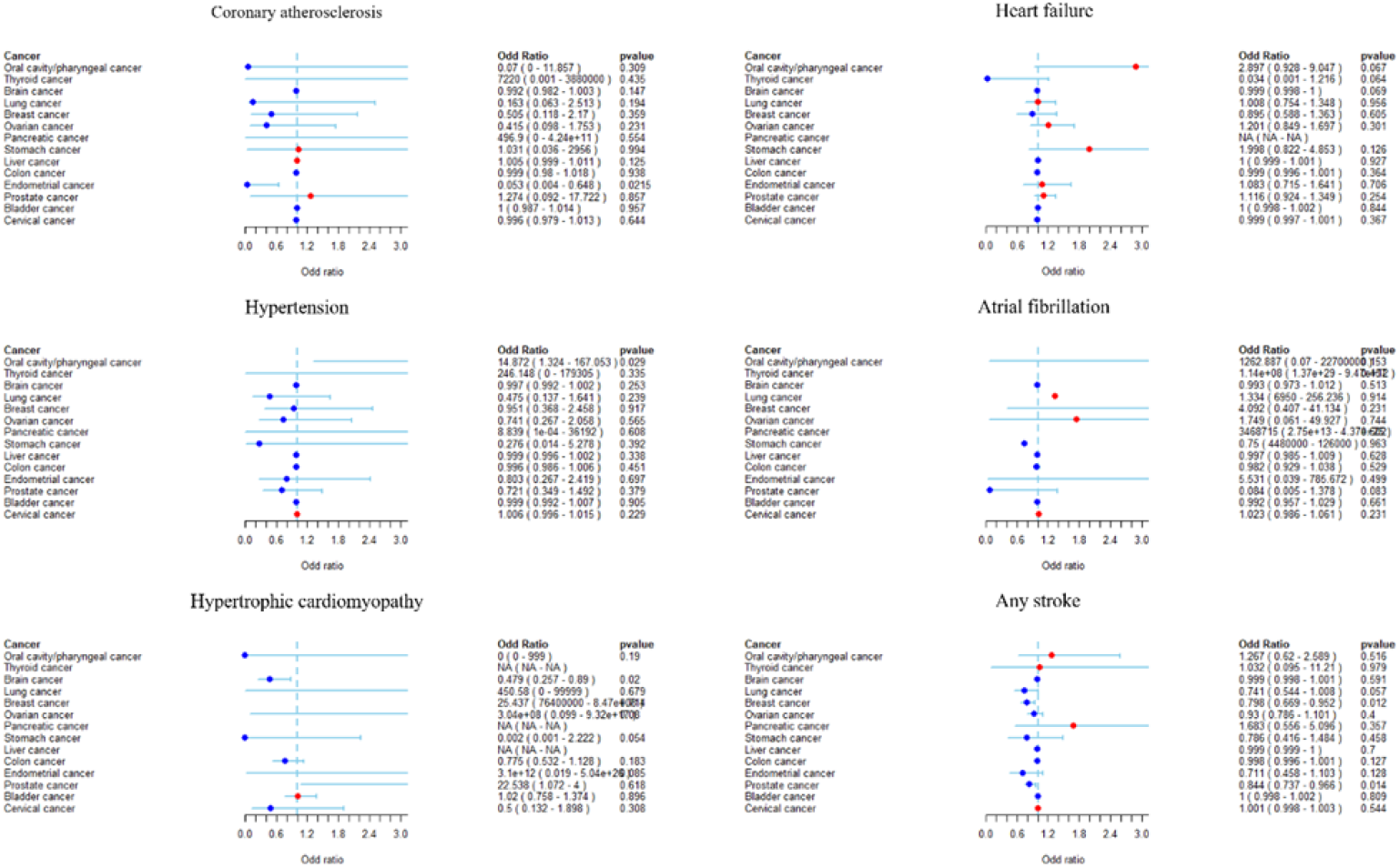
Associations of genetic liability to cardiovascular diseases with risk of cancers in the primary analysis.

### Causal Effects of Common Cancers on Coronary Atherosclerosis

Associations of genetic liability to cancer with the risk of coronary atherosclerosis were analyzed by the IVW method, and results of MR analyses are presented in **Figure 3** and **Supplementary Table S7**. Genetically predicted bladder cancer was significantly associated with an increased risk of coronary atherosclerosis (OR=1.426, 95% CI: 1.051-1.934, P= 0.023) and hypertension (OR=1.689, 95% CI: 1.115-2.557, P=0.013); pancreatic cancer was significantly associated with an increased risk of other strokes (OR= 1.047, 95% CI: 1.005-1.090, P= 0.027) but lost significance after multivariate testing. The following conclusions were stable after multivariate testing: prostate cancer was significantly associated with an increased risk of heart failure (OR= 1.030, 95% CI: 1.009-1.053, P= 0.006); cervical cancer was significantly associated with an increased risk of other strokes (OR= 8.751686e+03, 95% CI: 35.043-2.185650e+06, P= 0.001);

**Figure 3.**
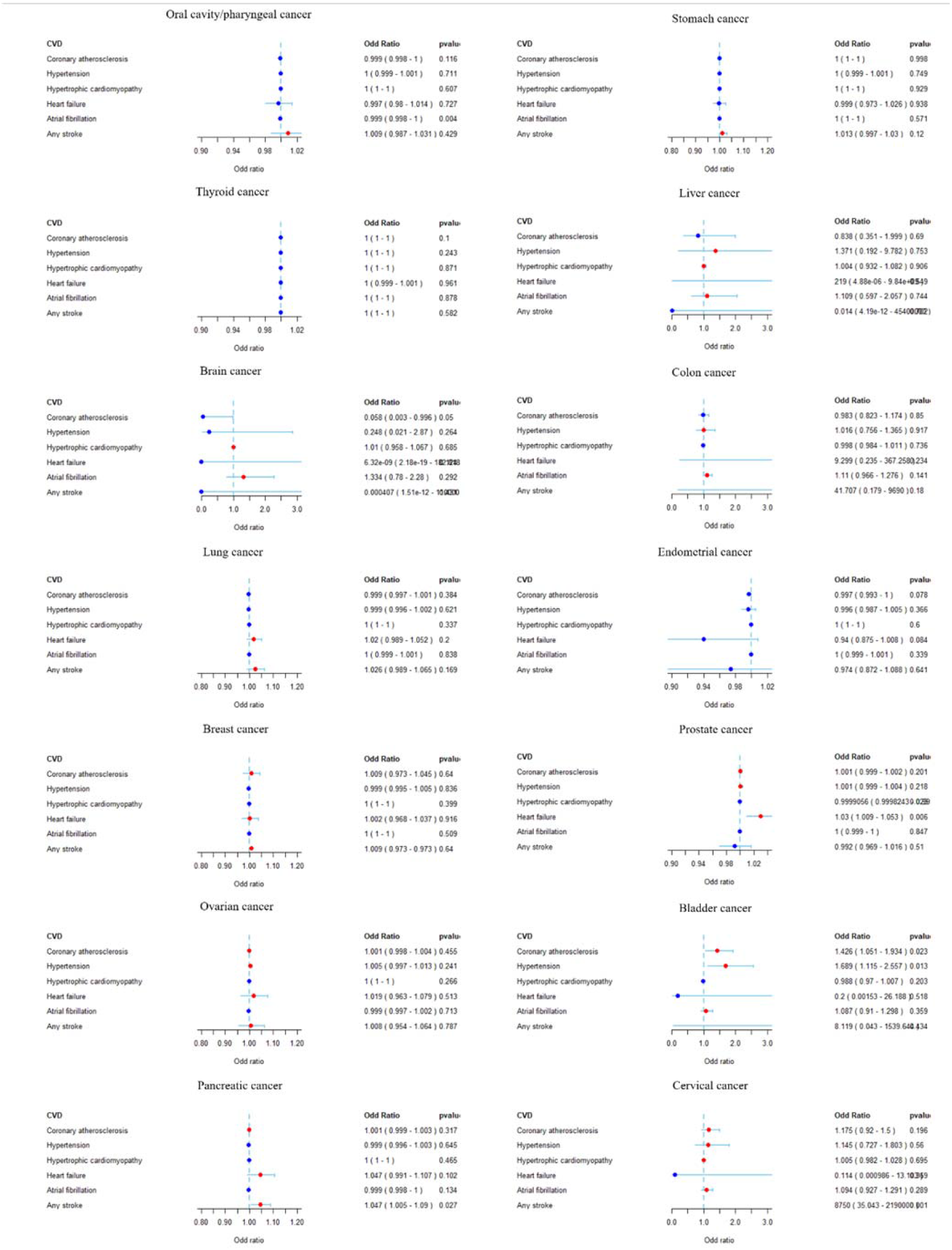
Associations of genetic liability to cancers with risk of cardiovascular diseases in the primary analysis.

No outlier was identified after MR-PRESSO analysis (**Table S9**). The results of heterogeneity analyses and horizontal pleiotropy tests are shown in **Tables S8** and **S10**. For the association between prostate cancer and heart failure, no heterogeneity among estimates of individual SNPs was detected in the Cochran Q-test (Q= 113.31; P= 0.154) and no pleiotropy was observed in the MR-Egger regression analysis (intercept P=0.591). The same characteristic was applied to an association between pancreatic cancer and other strokes and cervical cancer and other strokes. MR-Steiger analysis showed that the directions of causal associations were statistically significant (**Table S11**).

For the exposure factors in the above meaningful relationships, data from the corresponding FinnGen database were re-obtained and MR analysis was performed (**Table S12**). Coronary atherosclerosis was found to be significantly associated with an increased risk of endometrial cancer (OR= 1.1223, 95% CI: 1.004-1.254, P= 0.042), however, this result was contrary to that obtained from the IEU data. Bladder cancer was significantly associated with an increased risk of hypertension (OR= 1.002, 95% CI: 1.001-1.004, P= 0.009), the same as that obtained from the IEU data. Meta-analysis results are shown in **Table 1** after merging the association results from different databases.

**Table1.** Meta-analysis results after merging the association results from different databases.

| source | exposure | outcome | SNPs | P | OR | LCI | UIC |
| --- | --- | --- | --- | --- | --- | --- | --- |
| IEU | Coronary atherosclerosis | Endometrial cancer | 31 | **0.022** | 0.053 | 0.004 | 0.648 |
| finngen | Coronary atherosclerosis | Endometrial cancer | 40 | **0.042** | 1.122 | 1.004 | 1.254 |
| Combined |  |  |  | 0.449 | 0.321 | 0.017 | 6.092 |
| IEU | Hypertension | Oral cavity/pharyngeal cancer | 60 | **0.029** | 14.872 | 1.324 | 167.053 |
| finngen | Hypertension | Oral cavity/pharyngeal cancer | 107 | 0.075 | 0.834 | 0.683 | 1.019 |
| Combined |  |  |  | 0.112 | 0.851 | 0.697 | 1.038 |
| IEU | Hypertrophic cardiomyopathy | Brain cancer | 2 | **0.020** | 0.479 | 0.257 | 0.890 |
| finngen | Hypertrophic cardiomyopathy | Brain cancer | 3 | 0.411 | 1.000 | 1.000 | 1.001 |
| Combined |  |  |  | 0.413 | 1.000 | 1.000 | 1.001 |
| IEU | Any stroke | Breast cancer | 8 | **0.012** | 0.798 | 0.669 | 0.952 |
| finngen | Any stroke | Breast cancer | 11 | 0.647 | 0.989 | 0.943 | 1.037 |
| Combined |  |  |  | 0.274 | 0.975 | 0.931 | 1.021 |
| IEU | Any stroke | Prostate cancer | 8 | **0.014** | 0.844 | 0.737 | 0.966 |
| finngen | Any stroke | Prostate cancer | 11 | 0.782 | 1.009 | 0.948 | 1.073 |
| Combined |  |  |  | 0.439 | 0.978 | 0.924 | 1.035 |
| IEU | Bladder cancer | Coronary atherosclerosis | 34 | **0.023** | 1.426 | 1.051 | 1.934 |
| finngen | Bladder cancer | Coronary atherosclerosis | 23 | 0.075 | 1.001 | 1.000 | 1.002 |
| Combined |  |  |  | 0.074 | 1.001 | 1.000 | 1.002 |
| IEU | Bladder cancer | Hypertension | 35 | **0.013** | 1.689 | 1.115 | 2.557 |
| finngen | Bladder cancer | Hypertension | 25 | **0.009** | 1.002 | 1.001 | 1.004 |
| Combined |  |  |  | **0.009** | 1.003 | 1.001 | 1.004 |
| IEU | Prostate cancer | Heart failure | 101 | **0.006** | 1.030 | 1.009 | 1.053 |
| finngen | Prostate cancer | Heart failure | 31 | 0.386 | 0.989 | 0.963 | 1.015 |
| Combined |  |  |  | 0.123 | 1.013 | 0.997 | 1.030 |
| IEU | Pancreatic cancer | Any stroke | 6 | **0.027** | 1.047 | 1.005 | 1.090 |
| finngen | Pancreatic cancer | Any stroke | 14 | 0.887 | 1.002 | 0.971 | 1.035 |
| Combined |  |  |  | 0.138 | 1.019 | 0.994 | 1.045 |
| IEU | Cervical cancer | Any stroke | 21 | **0.001** | 8751.686 | 35.043 | 2185650.000 |
| finngen | Cervical cancer | Any stroke | 8 | 0.497 | 0.991 | 0.964 | 1.018 |
| Combined |  |  |  | 0.508 | 0.991 | 0.964 | 1.018 |

Abbreviation: SNPs, Single nucleotide polymorphisms; OR, Odds ratio; LCI, Lower confidence interval; UCI, Upper confidence interval.

## DISCUSSION

In our study, some potential causal relationships between CVD and cancer were found by bidirectional two-sample MR analysis. Hypertension increases the risk of oral cancer. Hypertrophic cardiomyopathy was significantly associated with a decreased risk of brain cancer. Stroke was significantly associated with a decreased risk of breast cancer and prostate cancer. Unfortunately, the significance of these causal associations reduced after adjustment for multiple testing, although their directionality was supported by the MR-Steiger analysis. For effects of common cancers on coronary atherosclerosis, we found that patients with bladder cancer were more likely to have coronary atherosclerosis and hypertension; pancreatic cancer increased the risk of stroke; these associations were attenuated after adjustment for multiple comparisons. The association of prostate cancer with increased risk for heart failure and for cervical cancer with increased risk for stroke remained robust after multiple testing. In the MR analysis of exposure factors obtained from the FinnGen database, an increased risk of bladder cancer for hypertension was also observed. MR analysis results suggest that partial genetic prediction of cancer subtypes and CAD subtype correlation warrant further study for the potential biological mechanism.

Several retrospective studies have investigated the effect of CVD on the risk of cancer.^28^ Framingham heart study (Framingham) and prevention of renal and vascular disease research (PREVEND) involving 20305 participants aged 36 to 64 years were assessed.^29^ The most common cancer subtypes during follow-up were those of the gastrointestinal (20%) tract, lungs (11%), prostate (16%), and breasts (18%). Those who maintained a heart-healthy lifestyle showed a reduced risk of CVD and a simultaneously lowered risk of cancer. An association between a heart-healthy lifestyle and lower cancer risk has been shown, and vice versa, a heart-unhealthy lifestyle is associated with higher cancer risk. Traditional CVD risk factors, such as age, male sex, smoking, or a history of smoking and diabetes are linked to increased risk of cancer, independent of the correlation. A retrospective cohort study with data derived from IBM’s MarketScan health insurance database^30^ showed that compared with patients without CVD, patients with CVD had a significantly higher risk of developing a new cancer. Sensitivity analysis was further performed with adjustment for smoking history and BMI, and the results remained significant. The results of subgroup analysis by tumor type suggested that the atherosclerotic CVD group could lead to an increased risk of lung cancer, bladder cancer, central nervous system cancer, and hematologic cancer. A Korean study reporting the association between hypertension and upper aerodigestive tract cancer found that patients with untreated hypertension showed the highest risk of developing oral cancer.^31^ This conclusion is consistent with the results of our study. Many similar retrospective studies have concluded that CVD is a high-risk factor for cancer development but it is interesting that hypertrophic cardiomyopathy is a protective factor for the brain and stroke was significantly associated with a decreased risk of breast cancer and prostate cancer as evidenced in the MR analysis. There are no studies on the effect of hypertrophic cardiomyopathy on the risk of cancer, and reports suggest that cancer therapy induces cardiomyopathy.^32^ Interestingly, an MR for women’s health study found an inverse association between coronary artery disease and breast cancer.^33^ Several factors suggested in the study may have contributed to these results. First, patterns may reflect ascertainment bias and competing risk. People at high risk of coronary heart disease (CHD) may live a shorter duration than those at low risk and are less likely to be diagnosed with breast cancer in their lifetime. Secondly, CAD treatment may be useful in preventing breast cancer. Third, the genetic mechanisms underlying CHD may have a protective effect on breast cancer. These explanations may also apply to the relationship of negative correlations in our study.

The phenomenon that cancer patients are more likely to suffer from CVDs is widely accepted by scientists but previous studies have found that this outcome is mainly caused by cardiovascular damage caused by anti-tumor treatment.^34^ The two diseases have common risk factors, such as old age, smoking, obesity, lack of exercise, etc.^35^ Cancer itself increases the risk of coronary thrombosis due to abnormal coagulation, platelet activation, and endothelial dysfunction.^36^ Recently patients with cancer and relative cardiac hypertrophy before receiving cancer treatment were studied^37^ and abnormalities in baseline cardiac function before antitumor therapy were reported.^38^ Therefore, cancer patients may be predisposed to CVD independent of the treatment. A study using data from the SEER database found that patients with breast cancer treated without chemotherapy or radiotherapy had a higher risk of developing a new CVD compared with the general population, especially when treated without tumor resection.^39^ This suggests that in addition to the side effects of radiotherapy and chemotherapy, genetic factors caused by cancer itself also have an impact on the subsequent occurrence of cardiovascular diseases. In a recent follow-up study of diagnosed UK Biobank participants, nearly one-third of cancer survivors had CVDs such as ischemic heart disease, stroke, arrhythmia (atrial fibrillation), and heart failure. The highest incidence of CVD was in patients with lung cancer (49.5%), blood cancer (48.5%), and prostate cancer (41%), and the risk of incident CVD was confirmed to be independent of traditional cardiovascular risk factors.^40^

This study has several strengths. First, it is the first systematic and comprehensive inclusion of cancer and CVD using a bidirectional MR analysis. Second is the MR study design, which alleviates confounding factors and reverses the causal bias. Our study yielded pooled data from large genetic studies in European populations, so the results are unlikely to be affected by population structure bias. Furthermore, high-quality data sources ensure adequate statistical power and robustness of the findings.

## STUDY LIMITATIONS

The limitations of the study should be considered. First, we cannot completely rule out the possibility that exposure to disease-associated SNPs affects the outcome of disease through other causal pathways, despite testing for horizontal pleiotropic effects and handling of outliers. Second, because this study only included Europeans, the generalizability of the findings is limited to other populations. Third, due to the use of summary data, we cannot draw nonlinear correlations or perform layered effect analysis. Our study was mainly based on available GWAS data. In the absence of GWAS for subtypes (pathological type, subsite, molecular subtype) in different cancers, it is difficult to infer the differential effect of further subtyping on causality. This study explored the relationship between cancer and CVD from a genetic perspective, and the mechanism different from previously determined common risk factors needs further investigation.

## CONCLUSIONS

In conclusion, this study is the first to analyze the causal relationship between common CVD and common cancer using a bidirectional MR method. We found several sets of associations as described above, although most of the conclusions are suggestive, especially that hypertrophic cardiomyopathy is a protective factor for brain and prostate cancer; it is contrary to previous findings. This study is expected to provide analysis ideas and directions for clinical management of chronic diseases and cancer.

## Data Availability

All data used in this study were obtained from GWAS summary statistics, which are publicly released by the Genetic consortium.

https://r6.finngen.fi/

https://gwas.mrcieu.ac.uk/

## ARTICLE INFORMATION

## Acknowledgments

This work was funded by the National Natural Science Foundation of China(31971166). We thank the included databases and the authors for generously making the data publicly available. We would like to thank KetengEdit (www.ketengedit.com) for its linguistic assistance during the language modification of this manuscript.

## Supplemental Material

Tables S1–S12

Figures S1–S2

## Contributors

Conception and design of the study: DMZ, XMX and YFZ; Collection of the data: HYL, YL, TT and DMZ; Analysis and interpretation of data; ZMF, YL, HYL and YFZ; Manuscript writing: DMZ, ZMF; Manuscript revising: XMX, YFZ and ZMF. Accesse and verify of the underlying data: TT, XMX.

